# Construction of a risk prediction model for postoperative bleeding in patients with thyroid cancer based on clinical data

**DOI:** 10.64898/2026.07.16.26358297

**Authors:** Weiyuan Chen, Xiaoxu Li, Wenqi Shen, Yan Zhang

**Affiliations:** Department of Thyroid and Breast Surgery, Affiliated Hospital of Xuzhou Medical University, Xuzhou 221000, China

**Author notes:** Corresponding author. (Z. Y). These authors contributed equally to this work (Weiyuan Chen, Xiaoxu Li).

**Keywords:** Thyroid cancer, Postoperative bleeding, Nomogram, Predictive model, Risk factors

## Abstract

**Objective:** To develop and validate a risk model for predicting postoperative bleeding in patients with thyroid cancer.

**Methods:** A total of 2800 consecutive patients diagnosed with thyroid cancer in the Department of Thyroid and Breast Surgery of the Affiliated Hospital of Xuzhou Medical University between January 2020 and December 2023 were retrospectively analyzed. Patients were categorized into two groups based on postoperative bleeding occurrence: bleeding and non-bleeding groups. Univariate and multivariate logistic regression analyses were utilized to screen independent risk factors. Meanwhile, risk prediction models were developed and nomogram. Subgroup analysis was performed to identify independent risk factors. The predictive effects of the models were assessed using the Hosmer-Lemeshow test and receiver operating characteristic (ROC) curves.

**Results:** Of the 2800 recruited patients, 50 had postoperative bleeding, with an incidence rate of 1.7%. Multivariate logistic regression analysis showed that age, hypertension, total thyroidectomy, tumor size ≥4 cm, and operation time ≥90 min were the risk factors for postoperative bleeding in thyroid cancer patients (P<0.05). A risk prediction model was established based on the above factors, and the area under the ROC curve was 0.881, with a sensitivity of 94.0%, a specificity of 67.3%, and an accuracy of 74.0%. Decision curve analysis revealed that the model had good predictive ability.

**Conclusions:** The constructed risk prediction model has good predictive power and can provide a reference for healthcare professionals to predict the risk of bleeding in patients after thyroid cancer surgery.

## 1. Introduction

Thyroid cancer is one of the most common head and neck malignancies. The estimated incidence rate of thyroid cancer in China was 14.6 per 100,000 persons in 2015, ranking seventh among all malignant tumors in China (National Health Commission of the People’s Republic of China Medical Administration and Hospital Administration, 2022). Surgical resection remains the mainstay of treatment for thyroid cancer (Lai et al., 2020), which can effectively resect the lesion, control tumor progression, and improve patient prognosis. Bleeding is one of the most serious early postoperative complications of thyroid cancer, occurring in 0.1-2.9% of postoperative thyroid cancer cases (Materazzi et al., 2017).

Despite its low incidence, it can cause asphyxia, hemiplegia, and even death if not properly managed (Liu et al., 2017). Therefore, identifying strategies to reduce the rate of postoperative bleeding in thyroid cancer has been the focus of clinical attention. While most of the current studies on postoperative hemorrhage in thyroid cancer focus on risk factor analysis, systematic research on constructing risk prediction models is largely lacking (Li et al., 2020). A nomogram line graph, also known as a vertical line graph, is a type of graph with scaled line segments drawn after integrating multiple risk factor indicators based on a multifactor regression model, which has the advantages of continuity, visualization, convenience and quickness in the prediction system, and can quickly assess the risk of clinical adverse events utilizing data visualization technology. It has been widely used in the field of medicine in recent years, and it can predict the incidence rate of the patient’s illnesses, the risk of progression, prognosis, etc. (Li et al., 2022). Although nomogramar plots are widely utilized in predictive models for various surgical complications, their application in constructing predictive models for postoperative bleeding in thyroid cancer has received less attention. Given its ability to assess the risk of an event based on indicators or characteristics that are closely related to the disease, a nomogram line graph has outstanding visualization advantages and is useful for guiding clinical nursing practice.

The present study sought to develop a risk model for predicting postoperative bleeding in patients with thyroid cancer to provide a clinical tool for the early identification of high-risk patients, optimize postoperative management, reduce complications, and provide a basis for the development of clinical care strategies.

## 2. Materials and methods

### 2.1 General information

Patients with thyroid cancer treated in the Department of Thyroid and Breast Surgery of the Affiliated Hospital of Xuzhou Medical University from January 2020 to December 2023 were retrospectively selected using a random sampling method. All patients underwent open thyroidectomy conducted by a specialized surgical team focused on thyroid procedures. During surgery, standard intraoperative management involved employing energy devices for both dissection and control of bleeding. Prior to closure, haemostasis was confirmed through careful inspection and, when necessary, the Valsalva maneuver. Drains were routinely inserted at the conclusion of the operation. Each case adhered to a standardized postoperative protocol, which included consistent monitoring of vital signs and thorough neck examinations.

Inclusion criteria: (1) patients aged ≥18 years old; (2) those meeting the diagnostic criteria of thyroid cancer (National Health Commission of the People’s Republic of China, 2019), confirmed by histopathology; (3) those with surgical indications for radical thyroid cancer surgery, thyroid cancer resection during hospitalization, undergoing the first operation, and operated by the same group of physicians; and (4) those with complete medical records and good compliance. Exclusion criteria: (1) patients with preoperative comorbidities of other primary malignant tumors; (2) those with severe organic diseases that could not tolerate surgery; (3) those with cognitive function and communication disorders; and (4) pregnant and lactating patients. The sample size was calculated using the dependent variable event count method (Events Per Variable, EPV) (Schmidt et al., 2015). EPV is calculated based on the number of events corresponding to each predictor. It is recommended by the statistical community that the value of EPV should be ≥10 to ensure the robustness of the multivariate logistic regression results. Referring to previous related studies, we predicted that up to five factors would be included in the final model (Qin et al., 2022). Using the criterion of an EPV of 10, we extrapolated the minimum need for an effective sample size to be 50 cases. Prior studies have shown that the rate of postoperative hemorrhage in thyroid cancer is about 0.1-2.9%; therefore, the estimated sample size was 1724, calculated as follows: (5 × 10) ÷ 2.9% = 1724. Considering a 10% shedding rate, 2800 cases were finally included in the analysis.

Given the low incidence of postoperative bleeding (1.7%), a case-control sampling approach was used for model development to ensure statistical stability. All 50 patients who experienced the bleeding group were included. From the remaining 2750 patients without bleeding, a random sample of 150 patients was selected to form the non-bleeding group,’ resulting in a final analysis dataset of 200 patients for the logistic regression analysis.

The study was approved by the Ethics Committee of the Affiliated Hospital of Xuzhou Medical University (XYFY2024-KL452-01).

### 2.2 Sampling Method and Grouping

Postoperative bleeding was defined as a clinically significant hematoma that necessitated surgical re-intervention for evacuation and haemostasis within 24 hours following the initial thyroidectomy. This endpoint was established based on the documentation provided by the attending surgeon in the medical records. Among the initial cohort of 2800 patients, 50 cases of postoperative bleeding were identified, yielding an incidence rate of 1.7%. To address the class imbalance and enhance statistical stability for model development, a case-control sampling approach was utilized. All 50 patients experiencing postoperative bleeding constituted the case group. From the remaining 2750 patients without bleeding, a random sample of 150 was selected using a computer-generated random number sequence to form the control group. This resulted in a final analysis dataset comprising 200 patients, with a case-to-control ratio of 1:3. The proportion of missing data for all variables was less than 1%. To address these missing values, multiple imputation was conducted using the Multiple Imputation by Chained Equations (MICE) method, with 5 imputations performed to ensure the reliability of model predictions.

### 2.3 Surgical and Perioperative Context

All patients underwent open thyroidectomy, carried out via a specialized thyroid surgical team.Standard intraoperative management involved the use of energy devices for dissection and hemostasis. Meticulous inspection and, when necessary, the Valsalva maneuver were employed to confirm hemostasis prior to wound closure. Drains were routinely placed at the conclusion of the procedure. A standardized postoperative protocol was adhered to for monitoring vital signs and conducting neck examinations in all cases.

### 2.4 Data Collection and Outcome Definition

Based on previous studies on factors associated with and predictors of postoperative bleeding in thyroid cancer (Zou and Yu, 2019), study nurses collected basic and disease-related information from patients undergoing surgery for thyroid cancer who met the inclusion criteria through the hospital’s electronic medical record system and nursing record searches. The collected data included gender, age, body mass index (BMI), comorbid hypertension, comorbid diabetes mellitus, mode of surgery, duration of surgery, tumor size, lymph node metastasis, amount of intraoperative bleeding, and postoperative cough. The primary outcome was postoperative bleeding. The outcome was defined as a clinically significant cervical hematoma and required reoperation within 24 hours after the initial thyroidectomy.Study nurses comprised three healthcare professionals from the thyroid surgery department and two master’s degree students in nursing who formed the investigative team. The survey team members were trained before conducting the survey, and after obtaining the consent of the head of the section, data were collected by the survey team members, and data collection was completed by April 2024. Data entry was verified by an associate chief nurse of thyroid surgery to ensure completeness, comprehensiveness, and accuracy of data collection. Multiple interpolation was used to estimate missing data, to ensure the reliability of the model predictions. The proportion of missing data for all variables was minimal (<1%). Multiple Imputation by Chained Equations (MICE) with 5 imputations was used to handle these missing values. To improve the accuracy of postoperative bleeding assessment and the study quality, each bleeding assessment was determined by the joint judgment of two research nurses with a bachelor’s degree or higher, ≥6 years of work experience, and previous experience in conducting nursing research.

### 2.5 Statistical analysis

Data were analyzed using SPSS 26.0 and R 4.1.2. Categorical count data were expressed as percentages and counts, and compared using the chi-square(X^2^) test. Variables with P<0.05 in the univariate analysis were included in the multivariate logistic regression analysis using the forward conditional method to identify independent risk factors. The results of the logistic regression are expressed as odds ratios (ORs) with 95% confidence intervals (CIs). The logistic regression equation was developed with the following reference categories: age (<60 years), hypertension (no), surgical procedure (partial resection), tumor size (<4 cm), and operation time (<90 min). A nomogram was constructed based on the results of the multivariate logistic regression model. The performance of the nomogram was evaluated in terms of discrimination and calibration. Discrimination was assessed by the area under the receiver operating characteristic (ROC) curve (AUC). Calibration was evaluated using calibration curves and the Hosmer-Lemeshow (H-L) goodness-of-fit test. The model was internally validated using the bootstrap method with 1000 repetitive samples to calculate a calibrated AUC and assess potential overfitting. The clinical utility of the model was assessed using decision curve analysis (DCA). A two-sided P<0.05 was considered statistically significant.

## 3. Results

### 3.1 Patient Characteristics

A comparison of the clinical data between the two patient groups is shown in Table 1. No statistically significant difference was found between the two groups regarding the proportion of patients with diabetes mellitus, lymph node metastasis, and Hashimoto’s thyroiditis and the number of postoperative coughs (P > 0.05). The proportion of patients who were males, aged ≥60 years, with BMI ≥24 kg/m2, hypertension, total thyroidectomy, tumor size ≥4 cm, hyperthyroidism, intraoperative bleeding ≥60 mL, and operation time ≥90 min was significantly higher in the hemorrhage group than in the non-hemorrhage group, and the difference was statistically significant (all P<0.05) (Table 1).

**Table 1.** Comparison of postoperative bleeding in 200 patients with thyroid cancer.

| Clinical information | Statistics<br>(*n* = 200) | Non-bleeding group<br>(*n* = 150) | Bleeding group<br>(*n* = 50) | $\chi^2$ | *P*-value |
| --- | --- | --- | --- | --- | --- |
| Genders |  |  |  | 5.546 | 0.019 |
| Male | 76 (38.0) | 50 (33.0) | 26 (52.0) |  |  |
| Female | 124 (62.0) | 100 (66.7) | 24 (48.0) |  |  |
| Age |  |  |  | 9.926 | 0.002 |
| <60 years | 136 (68.0) | 111 (74.0) | 25 (50.0) |  |  |
| ≥60 years | 64 (32.0) | 39 (26.0) | 25 (50.0) |  |  |
| BMI |  |  |  | 4.263 | 0.039 |
| <24 kg/m<sup>2</sup> | 85 (42.5) | 70 (46.7) | 15 (30.0) |  |  |
| ≥24 kg/m<sup>2</sup> | 115 (57.5) | 80 (53.3) | 35 (70.0) |  |  |
| High blood pressure |  |  |  | 12.923 | <0.001 |
| No | 130 (65.0) | 108 (72.0) | 22 (44.0) |  |  |
| Yes | 70 (35.0) | 42 (28.0) | 28 (56.0) |  |  |
| Diabetes |  |  |  | 0.800 | 0.371 |
| No | 157 (78.5) | 120 (80.0) | 37 (74.0) |  |  |
| Yes | 43 (21.5) | 30 (20.0) | 13 (26.0) |  |  |
| Surgical Procedures |  |  |  | 28.541 | <0.001 |
| Partial thyroidectomy | 127 (63.5) | 111 (74.0) | 16 (32.0) |  |  |
| Total thyroidectomy | 73 (36.5) | 39 (26.0) | 34 (68.0) |  |  |
| Tumor size |  |  |  | 31.658 | <0.001 |
| <4 cm | 154 (77.0) | 130 (86.7) | 24 (48.0) |  |  |
| ≥4 cm | 46 (23.0) | 20 (13.3) | 26 (52.0) |  |  |
| Lymphatic node metastasis |  |  |  | 1.132 | 0.287 |
| No | 107 (53.5) | 77 (51.3) | 30 (60.0) |  |  |
| Yes | 93 (46.5) | 73(48.7) | 20 (40.0) |  |  |
| Hyperthyroidism |  |  |  | 5.516 | 0.019 |
| No | 178 (89.0) | 138 (92.0) | 40 (80.0) |  |  |
| Yes | 22 (11.0) | 12 (8.0) | 10 (20.0) |  |  |
| Hashimoto's thyroiditis |  |  |  | 1.181 | 0.277 |
| No | 166 (83.0) | 127 (84.7) | 39 (78.0) |  |  |
| Yes | 34 (17.0) | 23 (15.3) | 11 (22.0) |  |  |
| Intraoperative bleeding |  |  |  | 8.267 | 0.004 |
| <60 mL | 111 (55.5) | 92 (61.3) | 19 (38.0) |  |  |
| ≥60 mL | 89 (44.5) | 58 (38.7) | 31 (62.0) |  |  |
| Surgical time |  |  |  | 12.788 | <0.001 |
| <90 min | 119 (59.5) | 100 (66.7) | 19 (38.0) |  |  |
| ≥90 min | 81 (40.5) | 50 (33.3) | 31 (62.0) |  |  |
| Postoperative cough |  |  |  | 0.837 | 0.360 |
| No | 119 (59.5) | 92 (61.3) | 27 (54.0) |  |  |
| Yes | 81 (40.5) | 58 (38.7) | 23 (46.0) |  |  |

### 3.2 Logistic Regression Analysis and Nomogram Construction

Postoperative bleeding occurrence in thyroid cancer patients was the dependent variable, and statistically significant variables in the univariate analysis, including gender, age, BMI, hypertension, surgical procedure, tumor size, hyperthyroidism, intraoperative bleeding, and surgical time, were independent variables. The risk of postoperative hemorrhage was 3.176 times higher in patients aged ≥60 years than in those aged <60 years. In addition, the risk of postoperative hemorrhage in hypertensive patients was 2.443 times higher than in non-hypertensive patients. The risk of postoperative hemorrhage was 4.419 times higher in patients with total thyroidectomy than in those with partial resection. Moreover, the risk of postoperative hemorrhage was 8.454 times higher in patients with tumor sizes of ≥4 cm than in those with tumor sizes of <4 cm The risk of postoperative hemorrhage was 3.604 times higher in patients with an operative time of ≥90 min than in those with an operative time of <90 min (Table 2).

**Table 2.** Multivariate logistic regression analysis of factors influencing postoperative bleeding occurrence in thyroid cancer patients.

| Considerations | B | SE | *Wald*$\chi^2$ | *P*-value | *OR*(95% *CI*) |
| --- | --- | --- | --- | --- | --- |
| Age | 1.156 | 0.442 | 6.842 | 0.009 | 3.176 (1.336-7.551) |
| High blood pressure | 0.893 | 0.422 | 4.483 | 0.034 | 2.443 (1.069-5.583) |
| Surgical Procedure | 1.486 | 0.414 | 12.910 | <0.001 | 4.419 (1.965-9.938) |
| Tumor size | 2.135 | 0.469 | 20.675 | <0.001 | 8.454 (3.369-21.215) |
| Surgical time | 1.282 | 0.422 | 9.250 | 0.002 | 3.604 (1.578-8.234) |
| Constant | -3.876 | 0.528 | 53.917 | <0.001 | - |

To determine whether the identified risk factors could predict value independently of the extent of surgery, we performed subgroup analyses (Table 3 and 4. Among patients who underwent partial thyroidectomy, the tumor size >4cm (OR=5.543,95%CI=1.848∼16.626) was found to be an independent predictor of postoperative bleeding. In the total thyroidectomy subgroup, the results indicated that tumor size>4cm (OR=22.379,95%CI =4.027-124.380), surgical time>90 min (OR=9.737,95%CI=2.297-41.270), and age ≥60years (OR=8.714,95%CI=1.839-41301) were independent predictors of postoperative bleeding.

**Table 3.** Logistic regression analysis of postoperative bleeding in partial thyroidectomy groups.

| Considerations | B | S.E | Wald | P-value | OR(95%CI) |
| --- | --- | --- | --- | --- | --- |
| Age | 0.673 | 0.548 | 1.504 | 0.220 | 1.959(0.669-5.740) |
| High bloodpressure | 0.700 | 0.539 | 1.686 | 0.194 | 2.014(0.700-5.797) |
| Tumor size | 1.713 | 0.560 | 9.339 | 0.002 | 5.543(1.848-16.626) |
| Surgical time | 0.715 | 0.534 | 1.791 | 0.181 | 2.043(0.717-5.819) |
| constant | -1.674 | 0.538 | 9.665 | 0.002 | - |

**Table 4.**
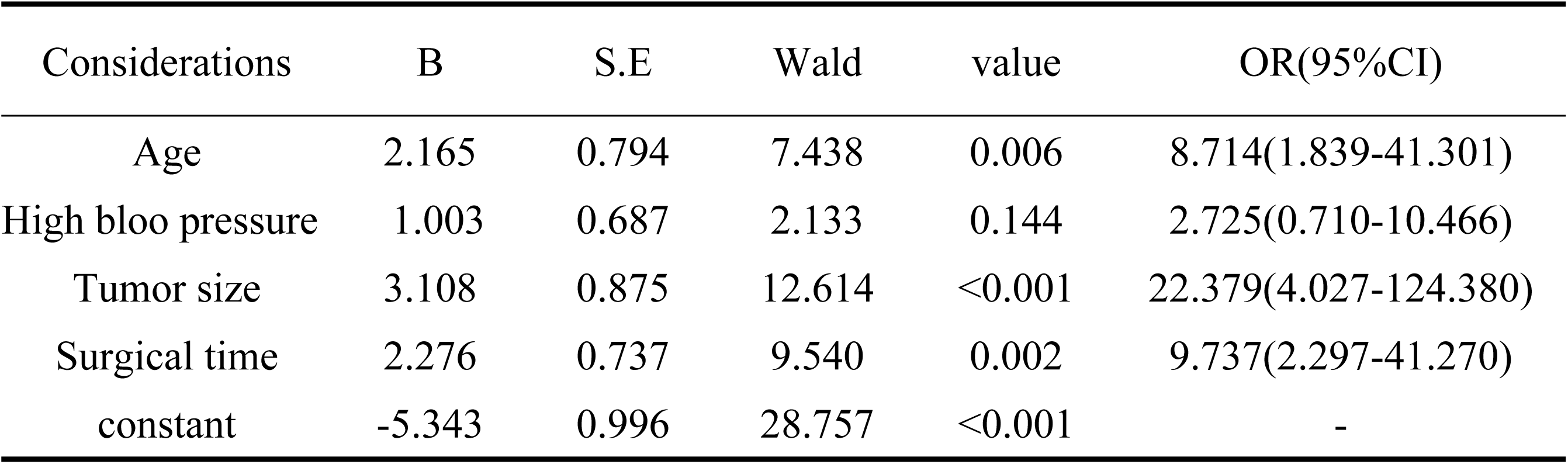
Logistic regression analysis of postoperative bleeding intotal thyroidectomy patients.

A risk prediction nomogram was constructed based on these factors (Fig. 1). The logistic regression equation was as follows: Ln(P/1 - P) = 1.156 × (Age: 1 if ≥60, 0 if <60) + 0.893 × (Hypertension: 1 if yes, 0 if no) + 1.486 × (Surgical Procedure: 1 if total thyroidectomy, 0 if partial) + 2.135 × (Tumor Size: 1 if ≥4 cm, 0 if <4 cm) + 1.282 × (Operation Time: 1 if ≥90 min, 0 if <90 min) - 3.876.

**Fig. 1.**
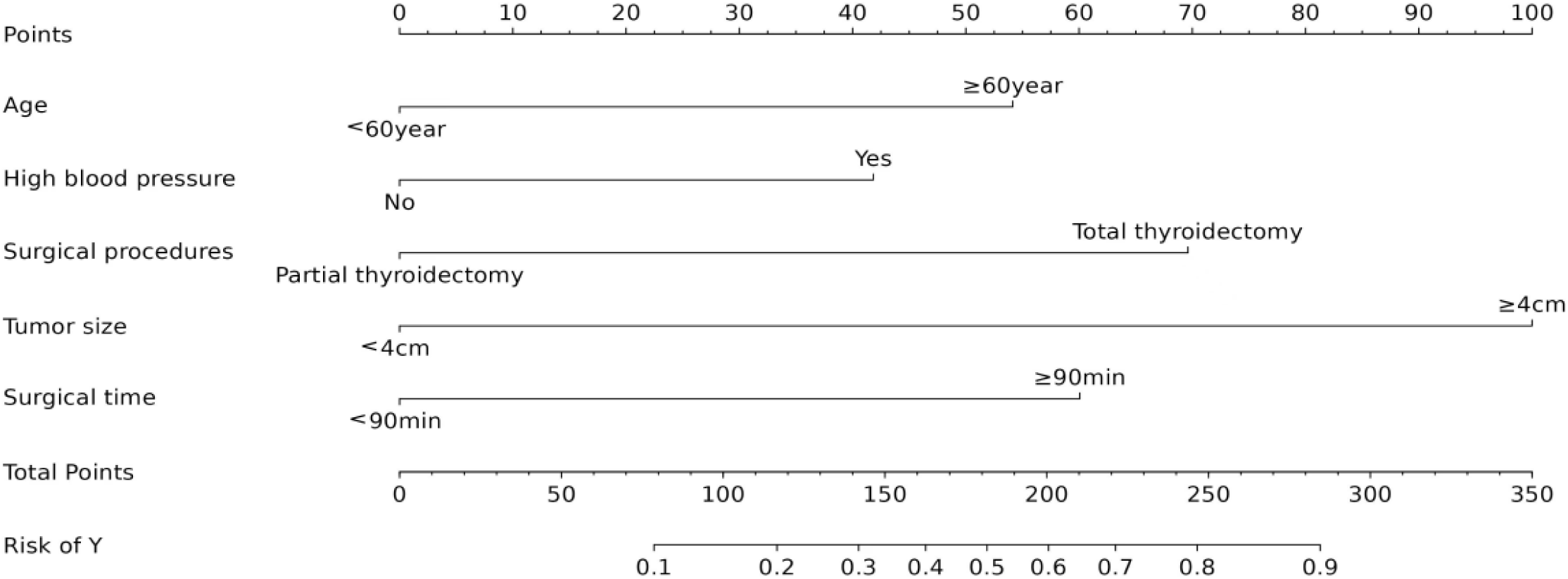
Multivariate logistic regression nomogram for predicting postoperative bleeding in patients with thyroid cancer.

### 3.3 ROC and calibration curve analyses

ROC and calibration curves of the nomogram are shown in Fig. 2. The area under the curve (AUC) was 0.881 (95% CI: 0.834-0.929), with a sensitivity of 94.0% and a specificity of 67.3%. Following internal validation with 1000 bootstrap samples, the calibrated AUC was 0.86. The Hosmer-Lemeshow goodness-of-fit test yielded a non-significant result (X² = 8.292, P = 0.308), and the calibration curve showed good agreement between predicted and observed probabilities (Fig. 3). The decision curve analysis (Fig. 4) demonstrated that the nomogram provided a net benefit across a wide range of threshold probabilities (0.03 to 0.7).

**Fig. 2.**
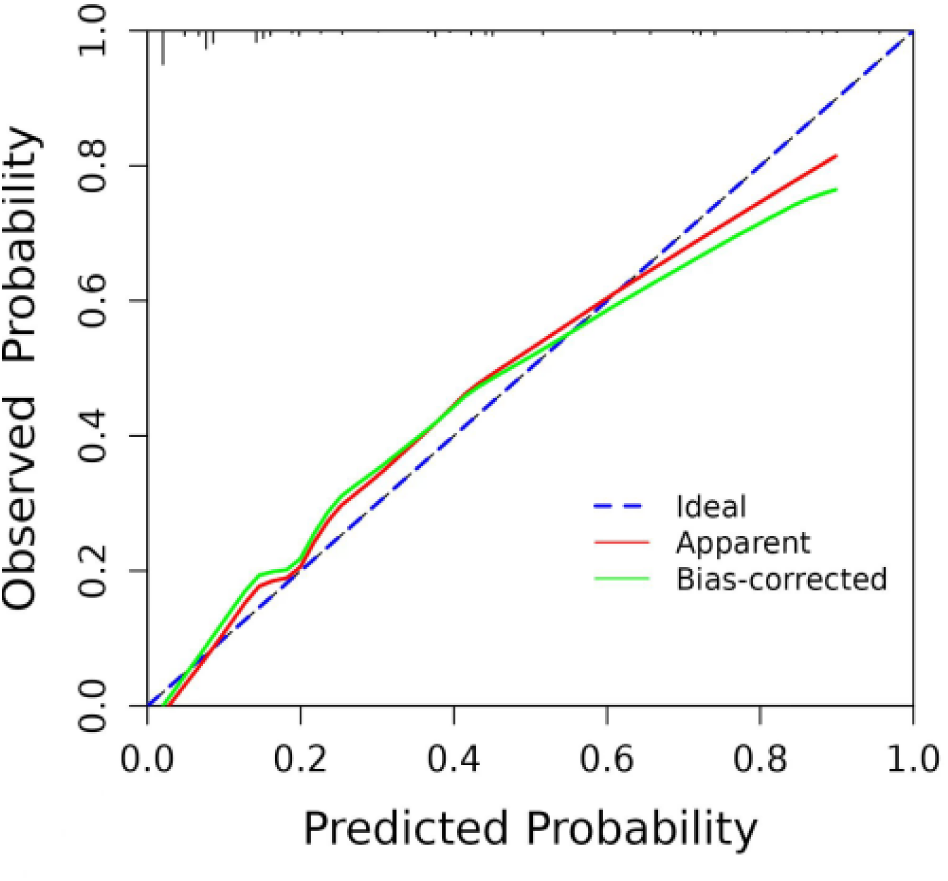
Validation of the nomogram model for predicting the risk of postoperative bleeding in patients with thyroid cancer.

**Fig. 3.**
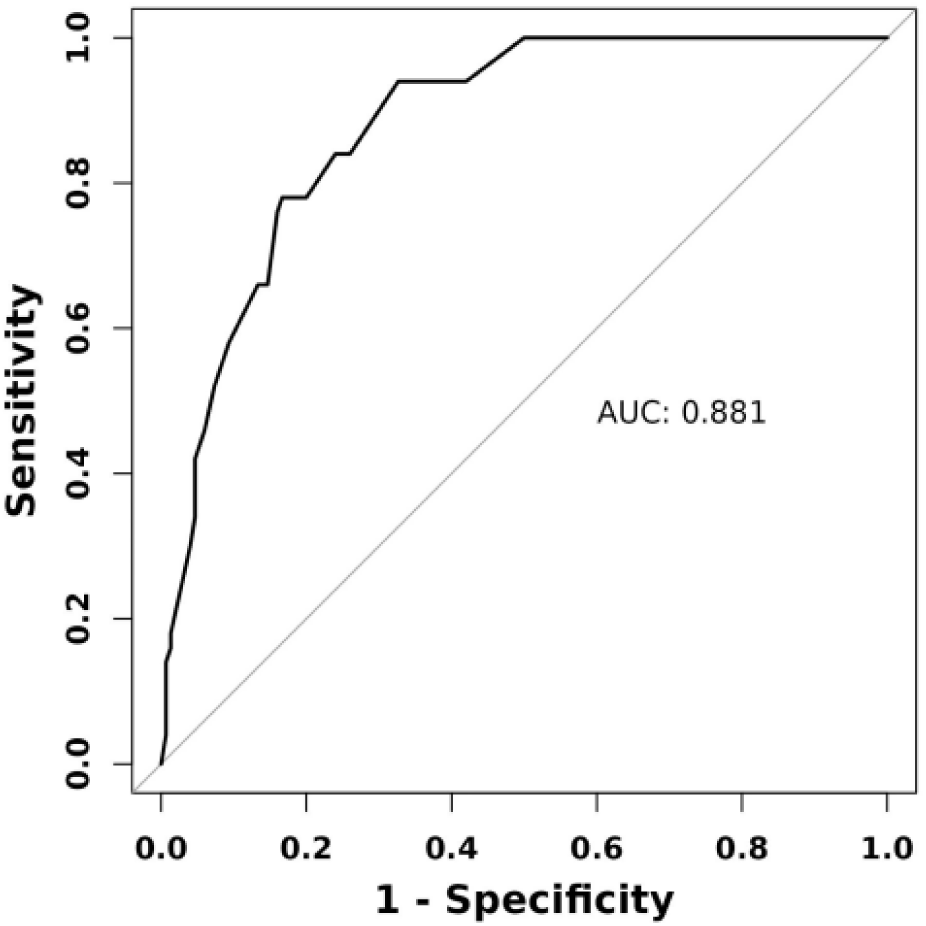
Nomogram prediction model of the risk of postoperative hemorrhage in thyroid cancer patients.

**Fig. 4.**
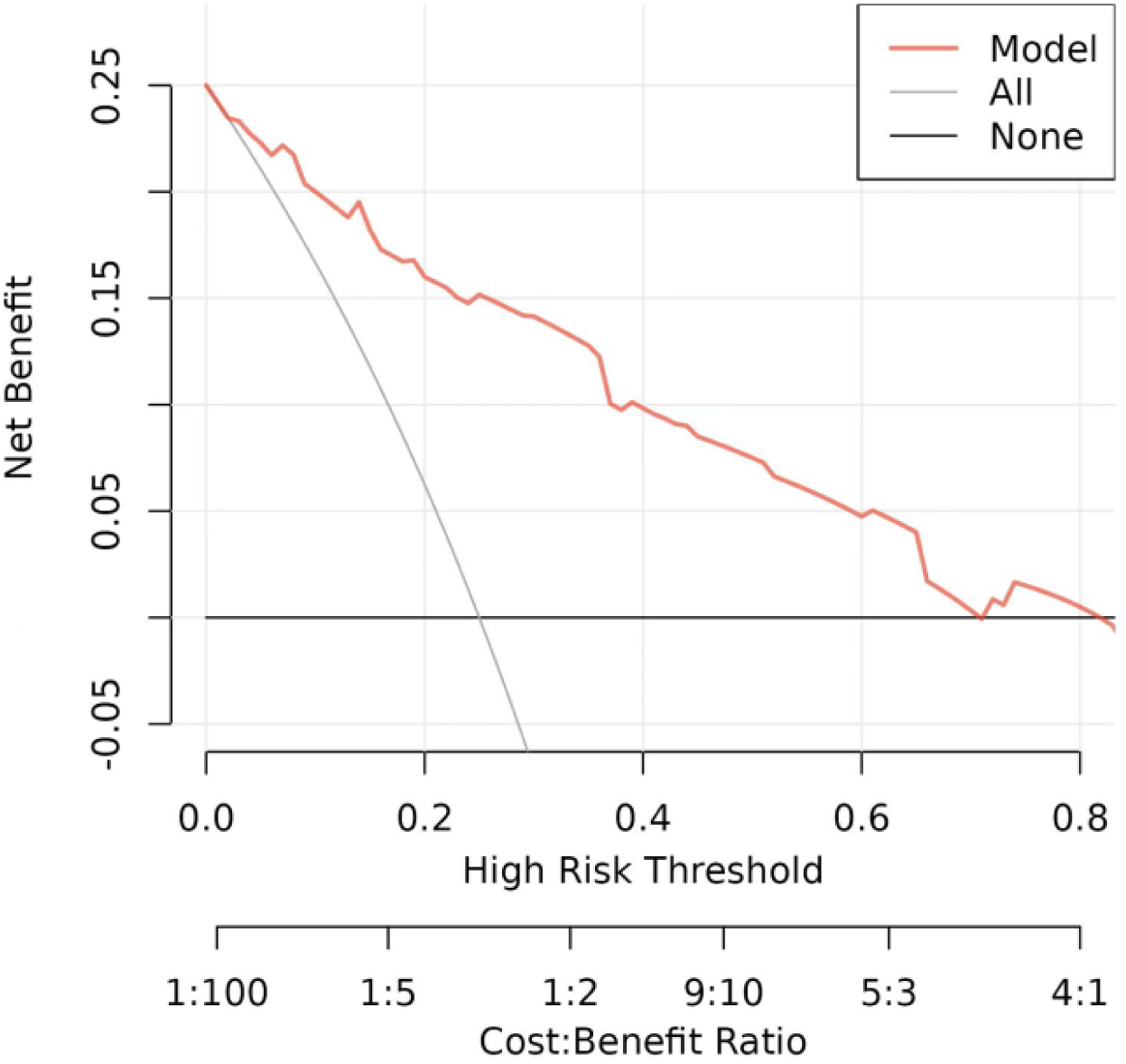
Decision curve analysis of the nomogram prediction model for the risk of occurrence of postoperative hemorrhage in patients with thyroid cancer.

Second, the constructed model was only internally validated using bootstrapping. External validation with a prospective, independent cohort from a different institution is recommended to confirm the generalizability and transportability of the proposed nomogram.. The results of the H-L goodness-of-fit test showed that the model is well differentiated (X^2^ = 8.292, P = 0.308>0.05).

## 4. Discussion

### 4.1 Summary of Main Findings

The incidence of thyroid cancer is increasing worldwide (He et al., 2024). Surgical resection is the first-line treatment for patients with thyroid cancer, resulting in an increased incidence of surgical complications such as hemorrhage, dyspnea and asphyxia, laryngeal nerve injury, and hypocalcemia (Pan et al., 2023). These subgroup analyses indicate that hypertension and prolonged operation time are significant risk factors, even within the less invasive partial thyroidectomy cohort. This finding implies that both vascular fragility and the complexity of surgery, as evidenced by extended operation durations, play a role in the risk of bleeding, independent of the extent of tissue resection. In contrast, patients undergoing the higher-risk total thyroidectomy exhibit a substantially increased risk when faced with large tumor sizes (≥4 cm), likely due to the heightened technical challenges and vascularity associated with such cases. This study successfully developed and internally validated a nomogram to predict postoperative bleeding in patients with thyroid cancer. The model integrates five readily available clinical variables: age, hypertension, surgical extent, tumor size, and operation time. The nomogram demonstrates strong discrimination, calibration, and clinical utility, offering a quantitative tool for personalized risk assessment.

### 4.2 Comparison with Existing Literature

The identified risk factors are consistent with the pathophysiological understanding of postoperative bleeding.Advanced age and hypertension are associated with decreased vascular elasticity and impaired coagulation (Lu et al., 2018). Total thyroidectomy and larger tumors (≥4 cm) entail a more extensive surgical dissection, increasing the potential vascular injury and technical difficulty (Li et al., 2020). A longer operation time (≥90 min) often reflects this increased complexity. While these factors have been reported previously, our study synthesizes them into a clinically actionable nomogram, which offers a significant advantage over considering factors in isolation.

### 4.3 Clinical Implications and Utility of the Nomogram

The primary value of this study lies in the creation of a practical nomogram. This tool can be integrated into perioperative care in several ways: Preoperative Planning: Blood pressure fluctuates greatly under the stimulation of intraoperative anesthesia and surgery, and venous return pressure rises further under the triggers of postoperative strenuous neck activities, coughing, and vomiting, thus increasing the likelihood of postoperative hemorrhage (Lu et al., 2018). Identifying high-risk patients allows for optimized preoperative preparation, such as meticulous blood pressure control in hypertensive patients and ensuring the availability of blood products.

Intraoperative Management: Postoperative hemorrhage can cause asphyxia, and if not properly treated in time, it can lead to hypoxic brain damage or even death (Yan et al., 2023). Thus, early diagnosis and rapid treatment are critical (Guo et al., 2018). Surgeons can exercise heightened vigilance regarding hemostasis in high-risk cases, potentially utilizing additional hemostatic techniques.

Postoperative Nursing Care: The model can guide risk stratification for nursing monitoring. Patients with a high nomogram score can be placed in more closely monitored settings, and nurses can be alerted to the early signs of hematoma formation, enabling faster intervention. Patient Education: Postoperative bleeding often increases the hospitalization time and hospital costs, reduces the quality of patient survival, and in severe cases can lead to asphyxiation or even death, causing medical disputes (Du et al., 2021). High-risk patients can receive more detailed counseling about warning signs, potentially leading to earlier presentation if complications arise.

### 4.4 Limitations and Strengths

This study has several limitations that must be acknowledged. First, its design as a single-center retrospective analysis may restrict the applicability of the findings to broader populations. To enhance the model’s transportability, external validation in prospective, multi-center cohorts is essential. Second, despite employing a case-control sampling strategy to ensure model stability, this approach may artificially inflate the event rate. The nomogram is calibrated to predict the odds of bleeding within this balanced dataset, necessitating recalibration for its use in the general population, where the incidence is only 1.7%. Third, the relatively small number of bleeding events (n=50) constrains the complexity of the model we could construct and may compromise the precision of our estimates. Fourth, while all procedures were conducted by a single surgical team to mitigate variability among surgeons, this choice limits the external validity of the findings; surgeon skill and volume remain potential unmeasured confounders. Nevertheless, a significant strength of this study lies in the rigorous development and internal validation of the model through bootstrap resampling, which yields a robust estimate of its performance.

## 5. Conclusion

In summary, the present study identified age, hypertension, total thyroidectomy, tumor size ≥4 cm, and operation time ≥90 min as the risk factors for postoperative bleeding in thyroid cancer patients. The nomogram model based on these factors had good differentiation and precision.

Moreover, this study constructed and validated for the first time a prediction model for postoperative bleeding in patients with thyroid cancer, which provides new evidence for the study of postoperative bleeding in thyroid cancer, and to some extent, provides a basis for the development of appropriate nursing strategies. However, this study has some limitations. First, this was a single-center, small-sample study. Second, although factors commonly associated with postoperative bleeding in thyroid cancer were included, this complication is associated with numerous factors, and some of the included factors, such as BMI, lymph node metastasis, and the amount of intraoperative bleeding, may not have been fully exploited, introducing a certain degree of bias. Finally, the established model was only validated internally, which, to a certain extent, limits the generalization and application of the study findings. Therefore, large-scale multicenter studies are warranted to construct accurate and reliable risk prediction models for postoperative bleeding in patients with thyroid cancer and validate them internally and externally to improve the generalizability and adaptability of the study.

## Declarations

## Ethics approval and consent to participate

Ethics committee approval was obtained fromthe Ethics Committee of the Affiliated Hospital of Xuzhou MedicalUniversity (decision number : XYFY2024-KL452-01).After ethics committee approval was obtained, participants were informed about the research on the planned data collection date and their informed consent was obtained.Participation in the study was voluntary.

## Consent for publication

Not Applicable.

## Data availability

All data reported in this study have been included in the manuscript.

## Author contributions

All the authors contributed to the study’s conception and design. Weiyuan Chenwrote the main manuscript text. Formal analysis began with Xiaoxu Li, and modifed as appropriate under the supervision of Yan Zhang. All the authors reviewed the manuscript.

## Declaration of interest

The authors report no declarations of interest.

## Acknowledgements

Our gratitude goes to all the women who consented to participate in this study and made the research worthwhile.

## Notes

### Competing Interest Statement

The authors have declared no competing interest.

